# Lesions in putative language and attention regions are linked to more severe strokes in patients with higher white matter hyperintensity burden

**DOI:** 10.1101/2021.11.05.21265496

**Authors:** Anna K. Bonkhoff, Sungmin Hong, Martin Bretzner, Markus D. Schirmer, Robert W. Regenhardt, E. Murat Arsava, Kathleen L. Donahue, Marco J. Nardin, Adrian V. Dalca, Anne-Katrin Giese, Mark R. Etherton, Brandon L. Hancock, Steven J. T. Mocking, Elissa C. McIntosh, John Attia, Oscar R. Benavente, John W. Cole, Amanda Donatti, Christoph J. Griessenauer, Laura Heitsch, Lukas Holmegaard, Katarina Jood, Jordi Jimenez-Conde, Steven J. Kittner, Robin Lemmens, Christopher R. Levi, Caitrin W. McDonough, James F. Meschia, Chia-Ling Phuah, Arndt Rolfs, Stefan Ropele, Jonathan Rosand, Jaume Roquer, Tatjana Rundek, Ralph L. Sacco, Reinhold Schmidt, Pankaj Sharma, Agnieszka Slowik, Martin Söderholm, Alessandro Sousa, Tara M. Stanne, Daniel Strbian, Turgut Tatlisumak, Vincent Thijs, Achala Vagal, Johan Wasselius, Daniel Woo, Ramin Zand, Patrick F. McArdle, Bradford B. Worrall, Christina Jern, Arne G. Lindgren, Jane Maguire, Polina Golland, Danilo Bzdok, Ona Wu, Natalia S. Rost, on behalf of the MRI-GENIE and GISCOME Investigators and the International Stroke Genetics Consortium

## Abstract

**Objective:** To examine whether high white matter hyperintensity (WMH) burden is associated with greater stroke severity and worse functional outcomes in lesion pattern-specific ways.

**Methods:** MR neuroimaging and National Institutes of Health Stroke Scale data at index stroke, as well as modified Rankin Scale (mRS) at 3-6 months post-stroke were obtained from MRI-GENIE study of acute ischemic stroke (AIS) patients. Individual WMH volume was automatically derived from FLAIR-images. Stroke lesions were automatically segmented from DWI-images, spatially normalized and parcellated into atlas-defined brain regions. Stroke lesion effects on AIS severity and unfavorable outcomes (mRS>2) were modeled within a purpose-built machine learning and Bayesian regression framework. In particular, interaction effects between stroke lesions and a high versus low WMH burden were integrated via hierarchical model structures. Models were adjusted for the covariates age, age^2^, sex, total DWI-lesion and WMH volumes, and comorbidities. Data were split into derivation and validation cohorts.

**Results:** A total of 928 AIS patients contributed to stroke severity analyses (mean age: 64.8(14.5), 40% women), 698 patients to functional outcome analyses (mean age: 65.9(14.7), 41% women). Individual stroke lesions were represented in five anatomically distinct left-hemispheric and five right-hemispheric lesion patterns. Across all patients, acute stroke severity was substantially explained by three of these patterns, that were particularly focused on bilateral subcortical and left-hemispherically pronounced cortical regions. In high WMH burden patients, two lesion patterns consistently emerged as more pronounced in case of stroke severity: the first pattern was centered on left-hemispheric insular, opercular and inferior frontal regions, while the second pattern combined right-hemispheric temporo-parietal regions. Bilateral subcortical regions were most relevant in explaining long term unfavorable outcome. No WMH-specific lesion patterns of functional outcomes were substantiated. However, a higher overall WMH burden was associated with higher odds of unfavorable outcomes.

**Conclusions:** Higher WMH burden increases stroke severity in case of stroke lesions involving left-hemispheric insular, opercular and inferior frontal regions (potentially linked to language functions) and right-hemispheric temporo-parietal regions (potentially linked to attention). These findings may contribute to augment stroke outcome predictions and motivate a WMH burden and stroke lesion pattern-specific clinical management of AIS patients.

## Introduction

A substantial fraction of adults experience a stroke in their lifetime.^1^ Many stroke patients experience persistent sequelae, which often severely affect their daily lives, as measured by disability-adjusted life years (DALYs).^2^ The accurate prediction of individual stroke outcomes is of particular interest, given that these predictions could inform healthcare professionals, as well as patients and their families about realistic expectations and guide the planning of rehabilitation phases.^3^ Previous outcome prediction efforts, amongst others collaborative ones enabling large clinical data sets, have certainly spurred progress in recent years.^4^ However, current stroke outcome predictions still tend to level off at performances short of immediate clinical relevance. An enhanced understanding of potential factors influencing post-stroke outcomes, that are not yet consistently integrated in current outcome models, could thus contribute to great improvements. Outcomes after stroke have already been shown to be associated with pre-stroke brain health,^5,6^ rendering the exploration of corresponding biomarkers particularly promising. White matter hyperintensity (WMH) burden, a neuroradiological marker of small vessel disease,^7,8^ could serve as a proxy of this pre-existing brain health, or brain frailty on the other end of the spectrum. WMH burden is associated with a multitude of cardiovascular risk factors, such as hypertension, diabetes mellitus, atrial fibrillation and smoking in both healthy and stroke populations.^9,10,11^ Moreover, WMH burden was shown to be linked to cognitive decline, impairment and dementia.^12,13^ Ultimately, WMH burden, as a hallmark of excessive neurodegeneration, may indicate *physiological*,^14^ as well as *pathological* brain aging.^11^ A growing body of literature additionally points towards deleterious effects of WHM burden on stroke outcomes. WMH burden and decreased white matter integrity negatively influence both early and long-term neurological outcomes, such as the National Institutes of Health Stroke Scale (NIHSS)-based stroke severity in the first few days after admission,^15^ as well as the modified Rankin Scale (mRS)-defined functional outcome six months post-stroke.^16,17^ Higher WMH burden is also linked to higher risks of recurrent stroke, post-stroke dementia and all-cause mortality after stroke.^18^

The effects of WMH burden are particularly well studied in case of language impairments and their recovery after stroke. Several studies have uncovered significant links between WMH burden and (chronic) aphasia severity,^19,20^ a more pronounced deterioration of language abilities in chronic aphasia^21^, and decreased efficacy of language treatment in chronic aphasia.^22^ Despite these apparent links between WMH burden and behavioral abilities, it is currently unknown, whether WMH burden increases vulnerability to all strokes, or only to specific lesion patterns. In line with Georgakis and colleagues, who recommend exploring the prognostic effect of WMHs in observational studies of ischemic stroke patients,^18^ the current study aimed to dissect how the specific lesion location may interact with the level of WMH burden in a stroke outcome-relevant way. In fact, we hypothesized to extract lesion pattern-specific WMH burden effects. More precisely, we expected to find that WMH burden would especially interact with lesion patterns involving putative language areas, given the just displayed links between WMH burden and language function in previous studies. Hence, we expected stroke symptoms to be more severe in situations of high WMH burden and left-hemispheric lesions affecting language areas. We employed state-of-the-art Bayesian hierarchical regression to model lesion pattern effects on acute stroke severity and long-term functional outcomes in individuals with high and low WMH burden. We developed models in a large multicenter cohort and subsequently validated them in a single-center cohort. The promise of our work is to enhance our neuroscientific understanding of how WMH burden is linked to stroke outcomes. These insights could then be leveraged to improve the prediction of stroke outcomes in the longer term and increase the efficacy of acute and chronic rehabilitative efforts post-stroke.

## Methods

### Stroke patient population

The present study capitalizes on data of 3,301 AIS patients assembled within the framework of the international, multi-site MRI–Genetics Interface Exploration (MRI-GENIE) study,^23^ that, in turn, build upon the infrastructure of the Stroke Genetics Network (SiGN, c.f., **supplementary materials** for details).^24^ We here considered all those MRI-GENIE AIS patients with available high-quality DWI-derived lesion segmentations,^25^ FLAIR-derived WMH burden,^26^ NIHSS-defined stroke severity and/or mRS data (n=1,107 patients in total). We performed complete case analyses and thus excluded patients if information on sociodemographic/clinical characteristics was missing (age, sex, comorbidities; c.f., **supplementary materials** for a sample size calculation). Patients gave written informed consent in accordance with the Declaration of Helsinki. The study protocol was approved by Massachusetts General Hospital’s Institutional Review Board (Protocol #: 2001P001186 and 2003P000836).

### Clinical and neuroimaging data

Sociodemographic and clinical data included information on age, sex, hypertension (HTN), coronary artery disease (CAD), diabetes mellitus (DM), atrial fibrillation (AF), history of smoking and prior stroke. Outcomes of interest were the acute NIHSS-based stroke severity, measured at index stroke, i.e. during the acute hospital stay (0-42, 0: no measured deficits, 42: maximum stroke severity) and the long term mRS score, obtained between day 60 to day 190 post-stroke, binarized to favorable (0-2) versus unfavorable (3-6) outcome.^27^

### Neuroimaging data, preprocessing and low-dimensional lesion embedding

Axial T2-FLAIR and DWI images were acquired between 2003 and 2011 with the majority of scans being obtained within 48 hours of hospital admission (median: 2 days, interquartile range 1-4 days, 61% in first 2 days, 91% in first week). Given the multi-site character of MRI-GENIE and neuroimaging acquisition in routine clinical practice, imaging parameters differed slightly between centers as outlined in detail in **supplementary materials**. WMH lesion volume was derived from FLAIR images by a previously developed, fully automated deep learning-based segmentation pipeline.^26^ In brief, the segmentation pipeline featured total brain extraction and intensity normalization as preprocessing steps. WMH lesions were then automatically segmented using concatenated convolutional neural networks, that were specifically designed for WMH lesions. Scans and segmentations were carefully quality controlled via automatic and manual routines (c.f., ^26^). Similarly, we automatically obtained DWI-based stroke lesion segmentations via an ensemble of 3-dimensional convolutional neural networks.^25^ DWI images and DWI-lesions were subsequently non-linearly normalized to MNI reference space and comprehensively quality controlled by two experienced raters (A.K.B, M.B.). Successively, the number of lesioned voxels within atlas-defined regions (94 cortical, 15 subcortical regions^28^ and 20 white matter tracts^29^) was computed. To further reduce the high dimensionality of the brain region- and white matter tract-space, we employed non-negative matrix factorization (NMF) to obtain ten unique spatial lesion patterns. The number of ten lesion pattern was chosen in line with our previous work^30,31^ and represents a trade-off between retaining as much lesion information as possible, while substantially reducing the lesion dimensionality.

### Identifying WMH-dependent brain substrates of acute stroke severity

The derived ten lesion patterns then represented the input of main interest to our Bayesian hierarchical regression framework.^32,33^ We aimed to infer the interaction effects of high and low WMH burden and lesion patterns in explaining stroke severity and functional outcomes. We, therefore, estimated relevances of lesion patterns separately for patients with high versus low WMH burden within our hierarchical model framework (c.f., **model specifications** in **supplementary materials**). We designated groups of patients as high and low WMH burden based on the median WMH burden after adjustment for patient age (resulting breakpoint: -5.3, c.f., **Table 1** for further characteristics, especially the median of WMH burden in high and low burden groups). To test the generalizability of our findings, we furthermore split the entire cohort into a multicenter derivation cohort and a single-center (Massachusetts General Hospital)^34^ validation cohort. Any of the regression analyses described in the following were thus performed separately in both cohorts for derivation and validation, respectively. We decided upon this derivation/validation splitting procedure, instead of bootstrapping or similar internal validation techniques, in view of the high computational burden of our Bayesian models.

**Table 1.** Clinical characteristics: Derivation cohort (acute stroke severity). We here present mean (SD) values, unless otherwise noted. We compared the groups of patients with low and high WMH burden via two-sample two-sample t-tests or two-sided Fisher’s exact test as appropriate. Asterisks indicate significant differences after family wise error-correction for multiple comparisons.

|  |  | **All participants**<br>**(n=664)** | **Low WMH burden**<br>**(n=332)** | **High WMH burden**<br>**(n=332)** | **Statistical comparison of patients with low and high WMH burden (FDR-corrected for multiple comparisons)** |
| --- | --- | --- | --- | --- | --- |
| **Age, years** | | 64.8 (14.5) | 63.6 (13.2) | 66.0 (15.7) | $p=0.08$ |
| **Sex (female)** | | 40.0% | 41.0% | 39.2% | $p=0.88$ |
| **NIHSS (median, interquartile range)** | | 4 (5) | 4 (5) | 4 (5) | $p=0.50$ |
| **Normalized DWI-derived stroke lesion volume (ml, median, interquartile range)** | | 3.1 (16.9) | 4.8 (25.6) | 2.3 (12.1) | $p=0.08$ |
| **Vascular lesion side** | **Left** | 43.8% | 43.1% | 44.6% | $p=0.90$ |
| | **Right** | 40.7% | 40.4% | 41.0% | $p=0.94$ |
| | **Both** | 7.5% | 8.1% | 6.9% | $p=0.88$ |
| **White matter hyperintensity lesion volume (ml, median,** |  | 5.77 (13.3) | 2.47 (3.03) | 15.35 (17.78) | **$p<<0.001^*$** |
| **interquartile range)** |  |  |  |  |  |
| **Cardiovascular risk factors** | **Hypertension** | 64.8% | 58.1% | 71.4% | ***p=0.002\**** |
|  | **Diabetes mellitus type 2** | 22.3% | 16.6% | 28.0% | *p=0.06* |
|  | **Atrial fibrillation** | 18.2% | 18.7% | 17.8% | *p=0.84* |
|  | **Coronary artery disease** | 17.9% | 16.0% | 19.9% | *p=0.39* |
|  | **Smoking** | 55.7% | 52.7% | 58.7% | *p=0.22* |
|  | **Prior stroke** | 6.8% | 3.6% | 9.9% | ***p=0.005\**** |
| **TOAST classification** | **Cardioembolic** | 23.3% | 23.8% | 22.9% | *p=0.90* |
|  | **Large artery sclerosis** | 16.4% | 19.3% | 13.6% | *p=0.12* |
|  | **Small vessel occlusion** | 14.9% | 19.3% | 18.4% | *p=0.06* |
|  | **Stroke of undetermined etiology** | 28.6% | 27.1% | 30.1% | *p=0.64* |
|  | **Stroke of other determined etiology** | 3.9% | 5.7% | 2.1% | *p=0.08* |

Next, we first aimed to explain the acute NIHSS-based stroke severity via hierarchical *linear* regression. Second, we explained unfavorable outcomes (mRS>2) via hierarchical *logistic* regression (we thus fitted regression models of stroke severity and unfavorable outcomes both in the derivation, as well as subsequently in the validation cohort). When modeling both of these outcomes, we accounted for several further covariates in addition to the lesion patterns: (mean-centered) age, age^2^, sex, smoking, hypertension, diabetes mellitus type 2, atrial fibrillation, coronary artery disease and prior stroke, as well as the log-transformed, *overall* stroke lesion volume and WMH lesion volume. All of these variables were determined a priori based on their conceivable associations to stroke severity and functional outcomes, as well as in line with our previous work.^30,31^ We included both age and age^2^ to carefully adjust for age-specific effects: By including both values, we could not only correct for linear, but also non-linear U-shaped age effects. Importantly, we also included the continuous WMH lesion volume to test for main effects of WMH burden in addition to interaction effects with stroke lesion patterns.

In accordance with the recommendations in Gellman and Hill (Chapter 3, p. 45-47),^32^ we tested the independence of errors assumption of the Bayesian linear regression model, and hence ensured the absence of any systematic violations of this assumption. We evaluated model performance via the coefficient of determination R-squared score (stroke severity model, linear regression) or the area under the curve (AUC, functional outcomes, logistic regression). We investigated differences in lesion pattern effects in patients with high and low WMH burden by comparing their posterior distributions (c.f., **WMH burden-specific lesion pattern effects** in **supplementary materials**). To decrease the likelihood of any biasing effects due to varying parcel-wise lesion volumes or frequencies of how often a parcel was affected by a lesion, we tested for differences between the groups of high and low WMH burden via independent two-sample t-tests or two-sided Fisher’s exact tests (level of significance: *p*<0.05, Bonferroni-corrected for multiple comparisons). In ancillary analyses, we repeated the described analysis workflow after assigning a low and high WMH burden status based on the median value of the raw WMH lesion volume, without initially regressing out an individual’s age.

### Data and code availability

The authors agree to make the data available to any researcher for the express purposes of reproducing the here presented results and with the explicit permission for data sharing by Massachusetts General Hospital’s and individual sites’ institutional review boards. The Harvard-Oxford and JHU DTI-based white matter atlases are accessible online (https://fsl.fmrib.ox.ac.uk/fsl/fslwiki/Atlases). Bayesian analyses were implemented in Python 3.7.

## Results

### Stroke patient population

A total of 1,107 patients were included in this complete case analysis study. The lesion data of all these 1,107 patients contributed to the derivation of the low-dimensional stroke representation. NIHSS and/or mRS scores were available for 931 out of these 1,107 patients and hence contributed to respective regression analyses. More precisely, we considered 928 patients in our stroke severity analyses (split into n=664 derivation, and n=264 validation, overall mean age: 65.0 (14.5), sex: 40% women) and 698 patients in our functional outcome analyses (split into n=542 derivation, and n=156 validation, overall mean age: 65.9 (14.7), sex: 41% women, c.f., **sample size calculation** in **supplementary materials**). The median NIHSS-based stroke severity at index stroke was 4 (interquartile range (iqr): 1-7) in the derivation and 3 (iqr: 0-6) in the validation cohort. With respect to long-term outcomes 27.9% in the derivation and 26.9% in the validation cohort experienced an unfavorable outcome (mRS>2). The WMH lesion volume was on average 5.77ml (median, iqr: 13.3ml) in the stroke severity derivation and 6.2ml (iqr: 13.5ml) in the stroke severity validation cohort. Further patients’ characteristics for the stroke severity derivation cohort are summarized in **Table 1**, an overlay of stroke lesions is presented in **Figure 1**.

**Figure 1.**
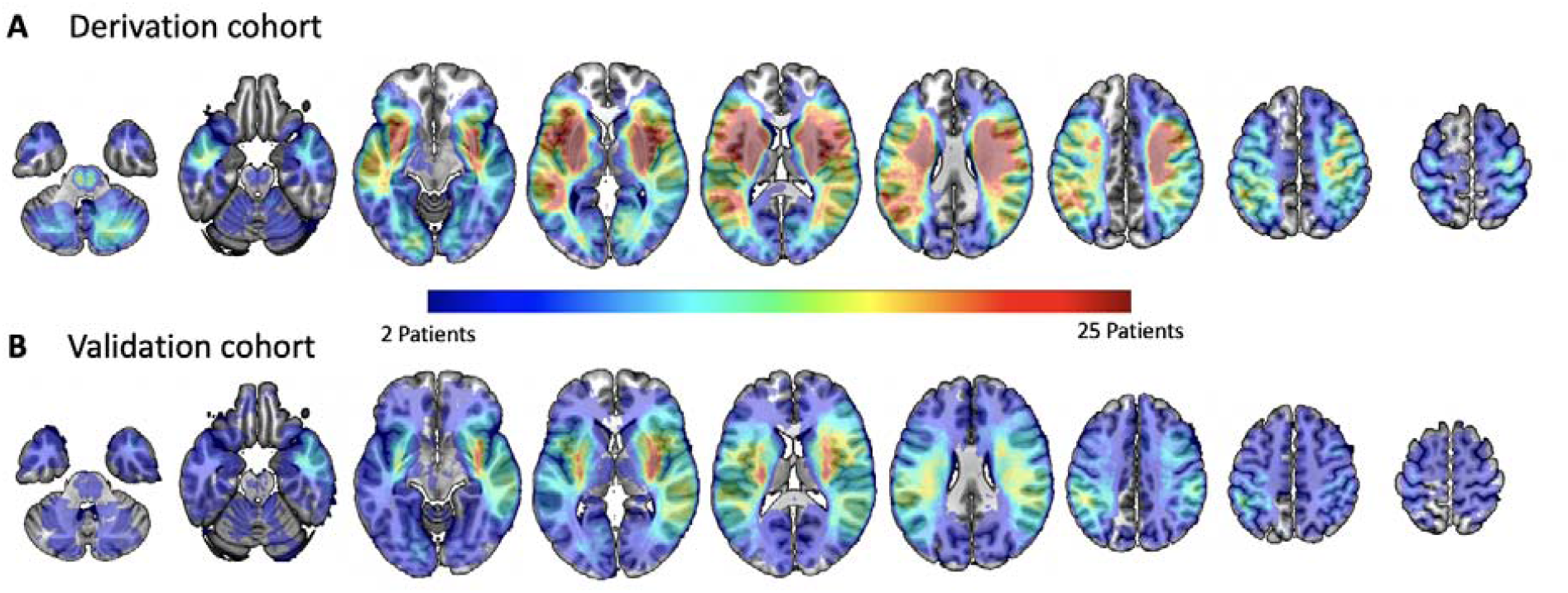
Lesions overlap of all patients in the stroke severity derivation cohort (A, n=664) and validation cohort (B, n=264). In both the derivation, as well as the validation cohort, the stroke lesion burden was evenly distributed between the left and right hemisphere with the maximal lesion overlap located bilaterally in subcortical brain regions.

### Anatomy of the extracted lesion patterns in stroke patients

We derived a low-dimensional stroke lesion representation via first computing lesion volumes per brain region and subsequent machine learning-based unsupervised dimensionality reduction. A high correlation between the original and reconstructed lesions indicated that important lesion information was retained despite dimensionality reduction (*r*=0.83, *p*<0.001). The resulting ten lesion patterns comprised anatomically plausible combinations of lesioned brain regions, equally distributed in the left and right hemispheres. Centers of these lesion patterns varied from anterior to posterior and from subcortical to cortical regions (**Figure 2**). Each patient’s individual lesion was represented by a combination of these ten lesion patterns.

**Figure 2.**
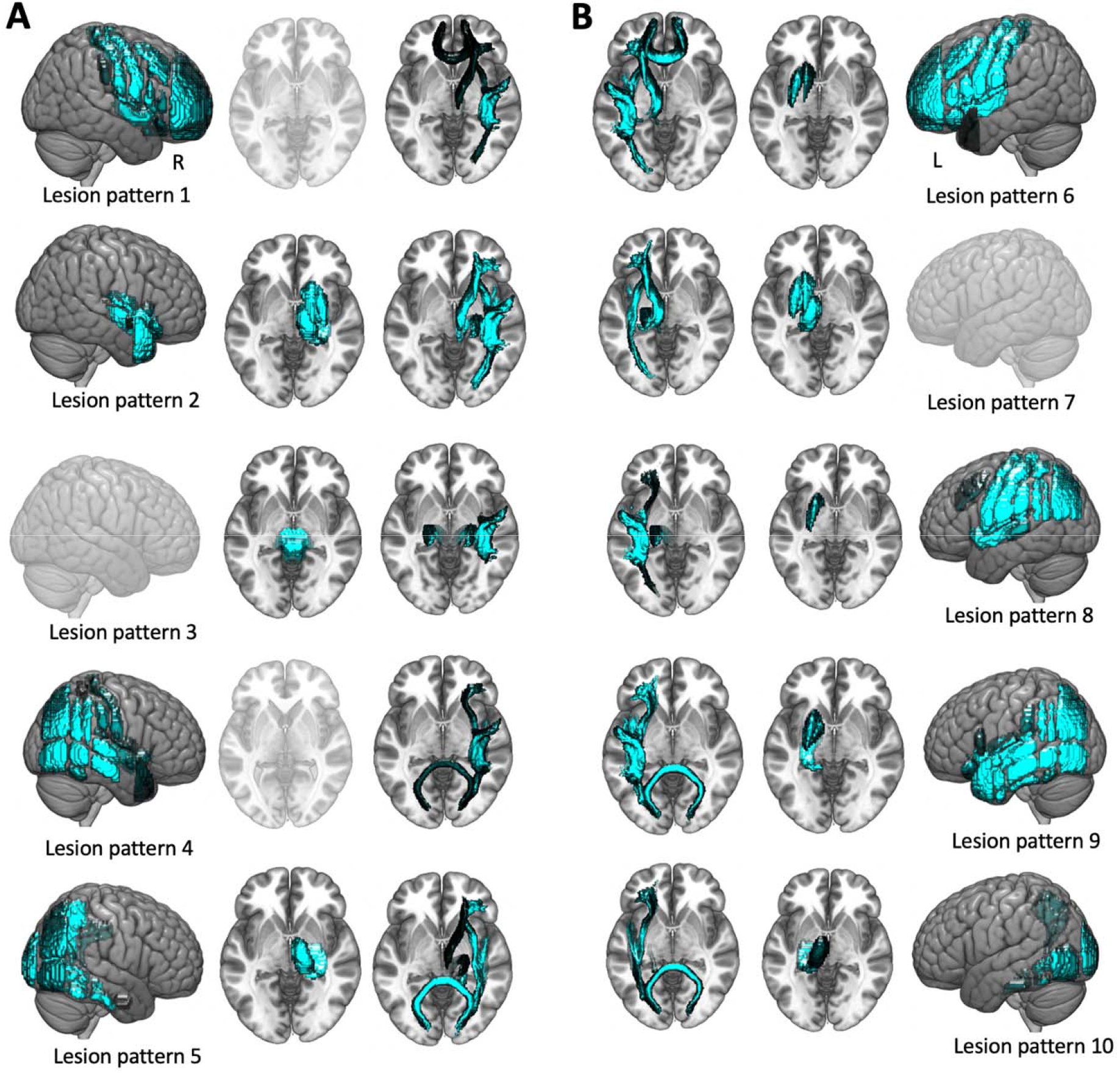
Individual stroke lesions captured in low dimensions via unsupervised machine learning techniques. Our automatically derived ten unique lesion patterns represented mainly right- (**A**) and left-hemispheric (**B**) strokes. The patterns featured anatomically plausible collections of brain regions with varying emphases ranging from cortical-subcortical and anterior-medial-posterior regions. Of note, there were five clearly left-hemispheric lesion patterns and four right-hemispheric ones. *Lesion pattern* 3 primarily comprised the brainstem, yet was nonetheless assigned to right-hemispheric strokes due to right-hemispherically more pronounced white matter tract contributions. Individual patients’ lesions were recreated by the weighted combination of lesion pattern. For example, a patient with a lesion in left subcortical brain regions would be characterized by high values for *lesion pattern* 7, but very low values for all other patterns. Renderings are shown in transparent in case of missing contributions from respective (sub)cortical regions.

### Explaining acute stroke severity

Successively, we aimed to explain acute stroke severity within the framework of our Bayesian hierarchical model, taking the ten lesion patterns as main inputs. Importantly, we modelled lesion pattern effects separately for patients with high and low WMH burden. WMH burden groups did not differ significantly with respect to the integrated covariates age, sex, stroke severity, stroke lesion volume, and most comorbidities, except for hypertension and prior stroke (hypertension: *p*=0.002, prior stroke: *p*=0.005, other covariates: *p*>0.05, Bonferroni-corrected for multiple comparisons, **Table 1**). Six lesion patterns (i.e., *lesion patterns* 1, 2, 3, 6, 7 and 8, c.f., **Figure 2 and 3**) substantially explained stroke severity across both high and low WMH burden groups. Lesion patterns comprising the brain stem, as well as bilateral subcortical regions had the highest mean posterior weights and were thus the most relevant in explaining stroke severity (*lesion patterns* 2, 3 and 7, **Figure 3**). With respect to individual brain regions, the highest weights were assigned to bi-hemispherical subcortical nuclei and white matter tracts, such as the anterior thalamic radiation, the corticospinal tract and inferior fronto-occipital and superior longitudinal fasciculus (**Figure 4A**). Cortical contributions were highest for the insula and opercular cortex and left-hemispherically pronounced pre- and postcentral cortex, inferior frontal, superior and middle temporal and supramarginal, as well as angular gyrus.

**Figure 3.**
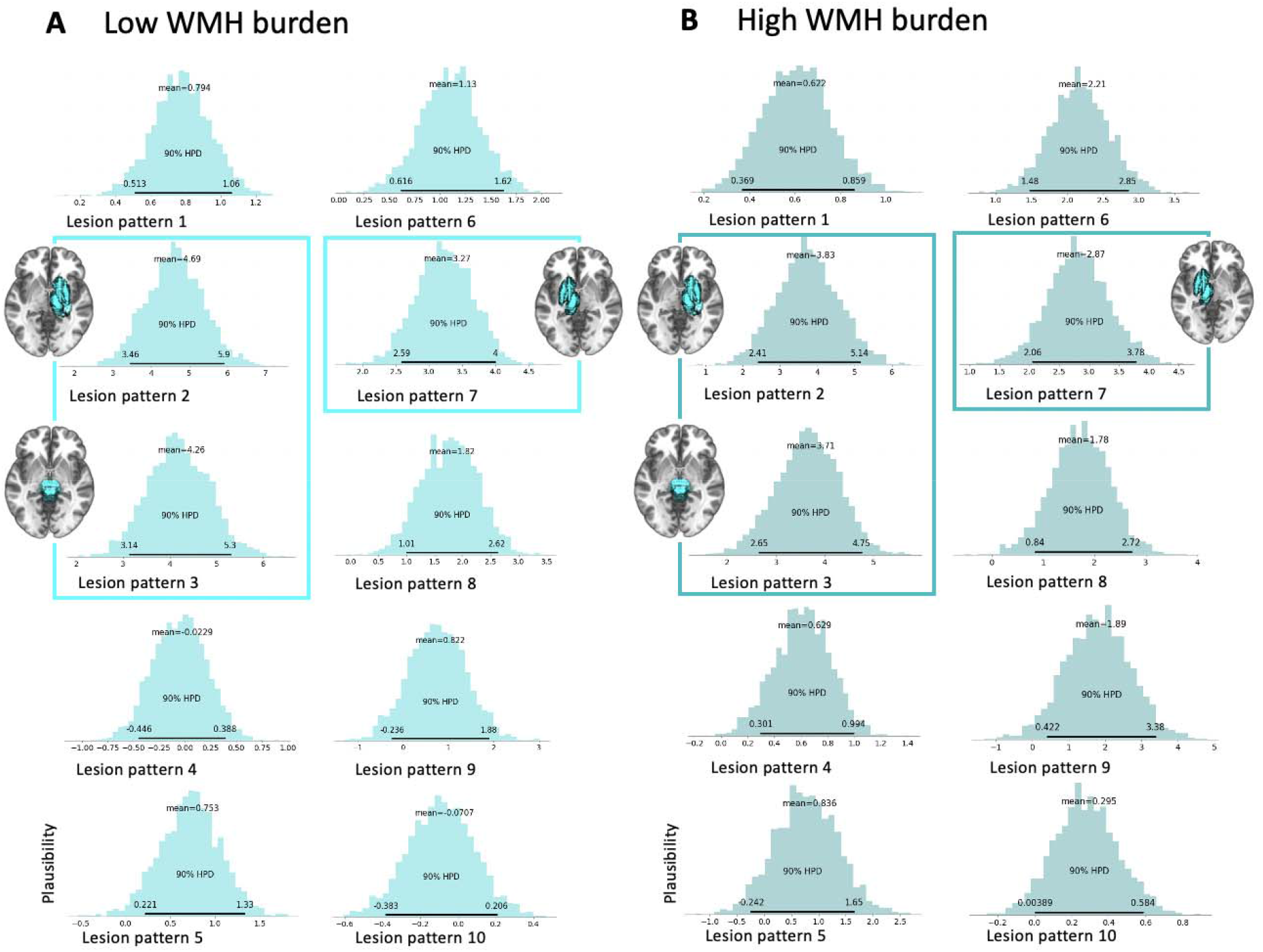
Bayesian posterior distributions indicating lesion pattern relevances in explaining stroke severity in patients with a low (A) and high (B) pre-stroke white matter hyperintensity burden. Six lesion patterns, *lesion patterns* 1, 2, 3, 6, 7 and 8, substantially explained stroke severity in both the low and the high WMH burden group, as indicated by the Bayesian posteriors distributions not substantially overlapping with zero (lower bound of the highest probability density interval (HPDI) of the posterior distribution covering 90%-certainty > 0). The three lesion patterns with the highest overall weights, *lesion patterns* 2, 3 of the right hemisphere and *lesion pattern* 7 of the left hemisphere, are specifically accentuated, i.e., framed, and accompanied by brain renderings of the respective lesion pattern (**low WMH burden:** *lesion pattern* 2: mean of the posterior distribution=4.7, 90%-HPDI=3.5 to 5.9; *lesion pattern* 3: mean=4.3, 90%-HPDI=3.1-5.3; *lesion pattern* 7: mean=3.3, 90%-HPDI=2.6 to 4.0; **high WMH burden:** *lesion pattern* 2: mean=3.8, 90%-HPDI =2.4 to 5.1; *lesion pattern* 3: mean=3.7, 90%-HPDI=2.7 to 4.8; *lesion pattern* 7: mean=2.9, 90%-HPDI=2.1 to 3.8). Together, these three patterns predominantly featured the brain stem, as well as bilateral subcortical grey matter regions (c.f., **Figure 4A**).

**Figure 4.**
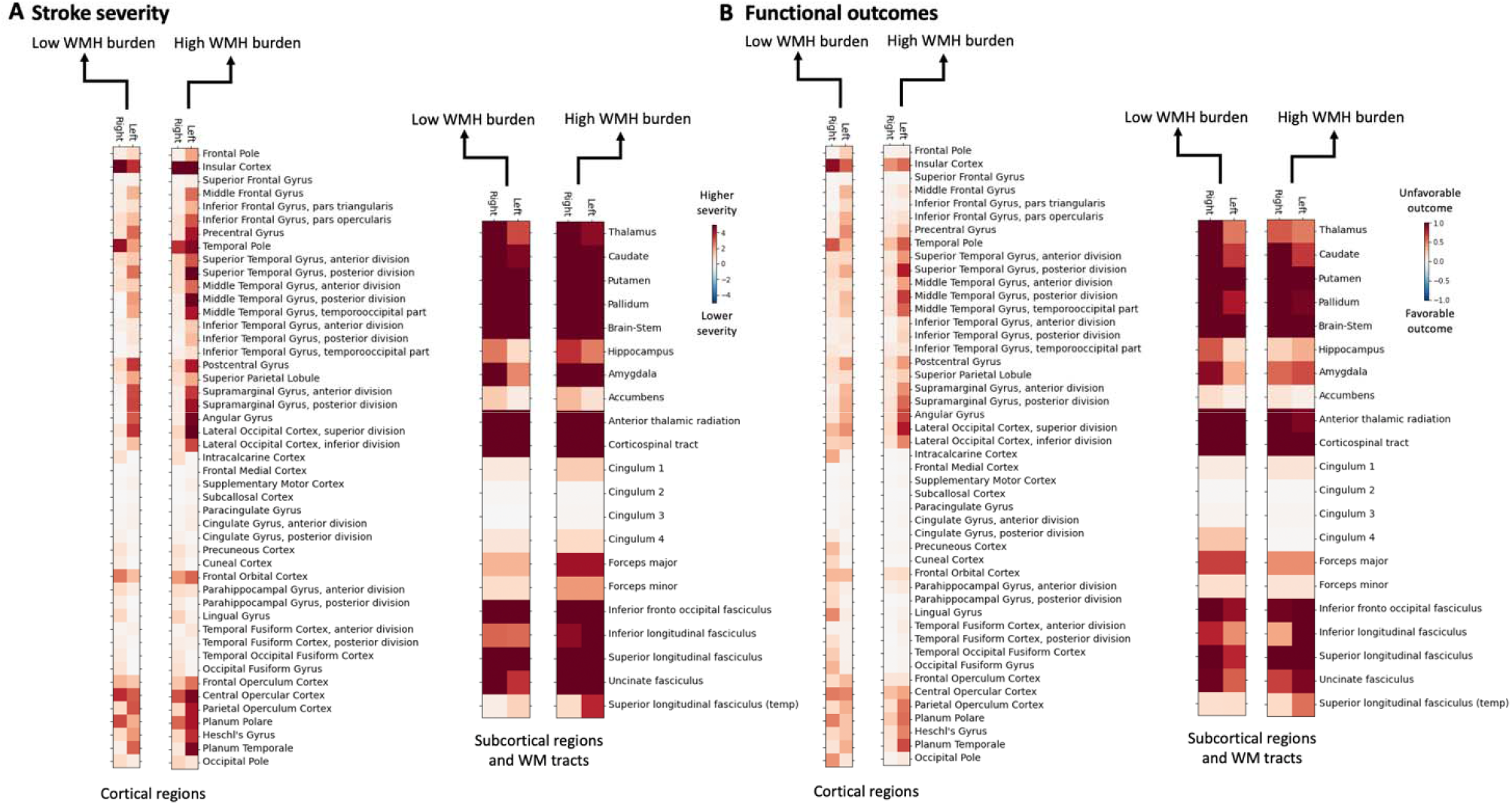
Brain region-wise relevances explaining NIHSS-based stroke severity (A) and mRS-defined functional outcomes (B) in the derivation cohort. Bilateral strokes affecting subcortical grey matter brain regions and white matter tracts explained higher stroke severity and higher odds of unfavorable outcomes in case of patients with both low and high WMH burden. In the case of stroke severity, cortical lesions had additionally relevant contributions that were, qualitatively and quantitatively (c.f., **Figure 5**), more pronounced for patients with a high WMH burden. The effect of cortical lesions was altogether weaker in the case of functional outcomes.

**Figure 5.**
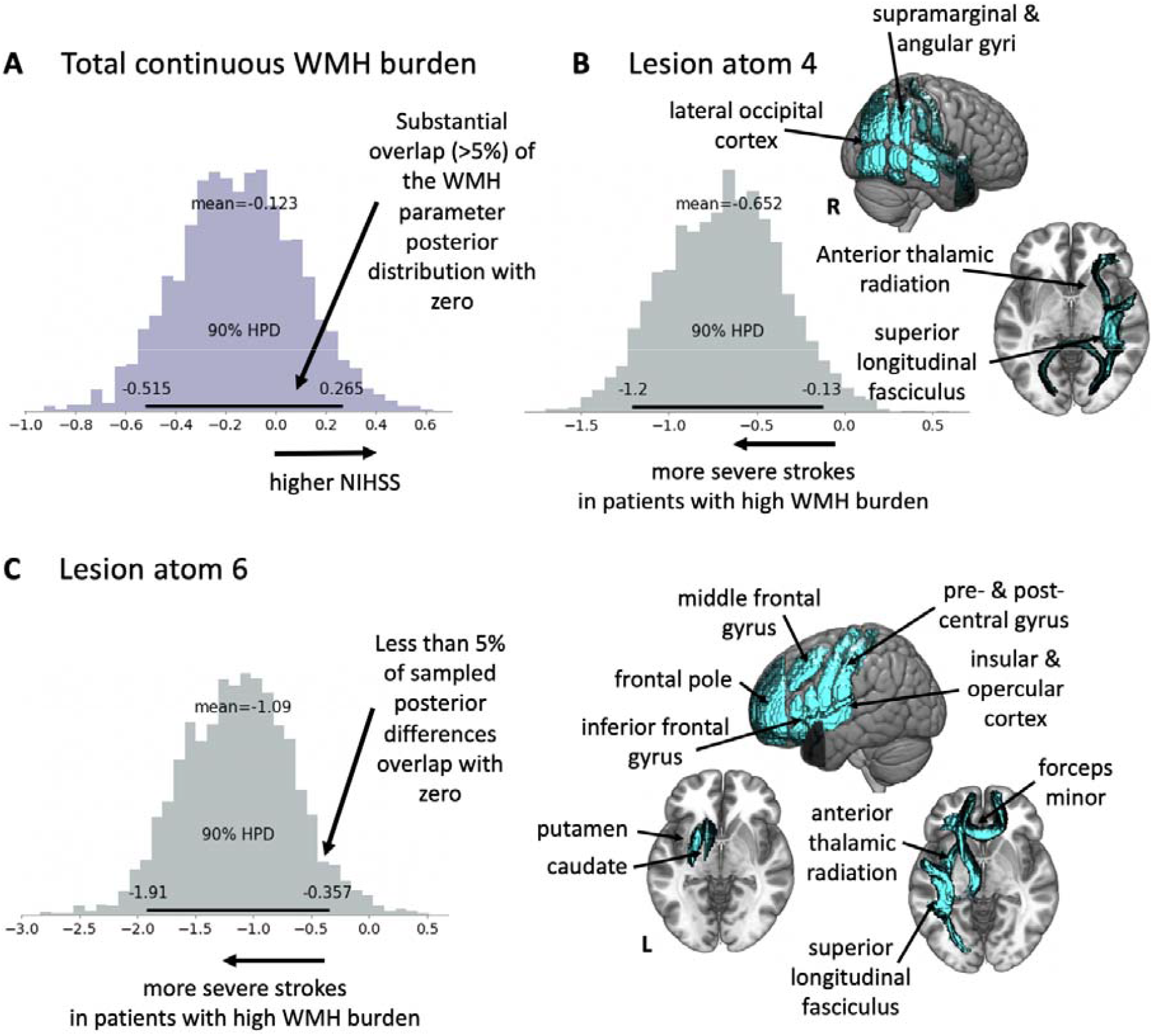
Substantial differences in lesion pattern effects for patients with low and high WMH burden. **A Bayesian posterior distribution of total WMH burden effect**. There was a substantial overlap (>5%) with zer for the Bayesian posterior distribution of the total WMH burden. Therefore, there was no evidence of an overarching total WMH burden effect on stroke severity. **B Substantially enhanced *lesion pattern* 4 effect in patients with high WMH burden**. We evaluated how much the Bayesian posterior distributions of coefficients for high and low WMH burden patients differed from one another by subtracting one posterior distribution from the other one, which resulted in difference distributions. In case of *lesion pattern* 4, a right-hemispheric lesion pattern, prominently featuring temporo-parietal brain regions, a higher stroke severity was specifically linked to those patients with a high WMH burden. This circumstance was inferable from the non-substantial overlap of the difference Bayesian posterior distribution with zero (*Lesion pattern* 4 (low – high WMH burden difference distribution): mean=-0.7, 90%-HPDI=-1.2 to -0.1). **C Substantially enhanced *lesion pattern* 6 effect in patients with a high WMH burden**. In addition, lesions of left-hemispheric brain regions, amongst others featuring the insular and opercular cortex, as well as inferior and middle frontal gyrus and pre- and post-central gyrus, were associated with a higher stroke severity in high WMH burden patients (*Lesion pattern* 6 (low – high WMH burden difference distribution): mean=-1.1, 90%-HPDI=-1.9 to -0.4).

We ascertained substantial differences between WMH groups in two lesion patterns. A lesion pattern in the left hemisphere, combining insular and opercular, as well as inferior frontal brain regions and a second right-hemispheric lesion pattern focused on temporo-parietal brain regions were substantially more relevant in high WMH burden patients than low burden (**Figure 5B & C**). In contrast, the overall, continuous WMH burden did not possess any additional substantial explanatory relevance (**Figure 5A**).

To give illustrative examples: A patient with a lesion affecting right temporo-parietal brain regions would be predicted to have a substantially higher stroke severity, if he or she had higher pre-stroke WMH burden, even if all other characteristics (age, sex, comorbidities etc.) were kept unchanged. The same could be said about a patient featuring lesions to left insular and opercular, as well as inferior frontal brain regions. Predicted stroke severity would be substantially higher in case of high WMH burden, even if nothing else about lesion or clinical characteristics was changed. However, in case of lesions primarily affecting brain regions other than the two named ones, the predicted stroke severity score would be the same independent of the WMH burden.

Similar results emerged when repeating the regression analysis of stroke severity in our validation cohort. In particular, the same two lesion patterns relating to left insular, opercular and inferior frontal and right temporo-parietal brain regions were more relevant in high WMH burden patients (c.f., **supplementary materials** for details).

### Explaining unfavorable functional outcomes

Our second aim was to investigate the effect of WMH burden on functional outcomes on average three months after stroke. We thus modeled favorable versus unfavorable functional outcome (cut-off: mRS>2) via Bayesian logistic regression, again disentangling effects for groups of high and low WMH burden by separately estimating their lesion pattern effects. None of the individual ten lesion patterns substantially explained unfavorable outcomes, as their posterior distributions rarely substantially deviated from zero (exception: *lesion pattern 4* for the high WMH group). Bilateral subcortical grey matter regions and white matter tracts were the most relevant when interpreting individual brain regions. Cortical regions were overall not as strongly implicated as in stroke severity, yet showed a left-hemispheric predominance, especially in the higher WMH burden group (**Figure 4B**). Furthermore, there were no substantial differences in lesion pattern effects between the high and low WMH burden groups. However, the total continuous amount of total WMH burden was associated with higher odds of unfavorable outcomes independent of stroke lesion locations.

More intuitively, our findings implied that a patient with a higher WMH burden had higher odds of an unfavorable outcome, independent of the location of the actual stroke lesion.

Results in our validation cohort were broadly similar: Subcortical regions in both hemispheres, as well as cortical regions in the left hemisphere were the most relevant ones. However, the overall effect of total WMH burden could not be replicated (c.f., **supplementary materials** for details).

### Ancillary analyses

In our main analyses, we accounted for the effect of aging on the WMH burden when defining the low and high WMH burden groups. In ancillary analyses, we defined low and high WMH groups based on the median of the *raw* WMH burden (breakpoint: 5.77ml). This breakpoint resulted in pronounced and significant age differences between the WMH burden groups (stroke severity: low WMH burden group: mean age: 58.2 (15.1) years, high WMH burden group: mean age: 71.3 (10.4) years, *p*-value<0.001; functional outcomes: low WMH burden group: mean age: 58.8 (15.3) years, high WMH burden group: mean age: 71.6 (9.8) years, *p*-value<0.001). Bayesian hierarchical model results, however, remained broadly similar. When modeling stroke severity, *lesion pattern* 6 was the most reliably emerging one: Patients with a high WMH burden were characterized by a substantially higher relevance of *lesion pattern 6* than those with a low WMH burden. In contrast, the substantial difference in *lesion pattern* 4 was not ascertainable anymore. In case of functional outcomes, the relevance of the overall continuous raw WMH burden remained apparent in the derivation cohort and was not present in the validation cohort, alike in the main derivation analyses.

There were no widespread, systematic differences in the frequencies of how often each cortical and subcortical grey matter region or white matter tract was affected, or in the numbers of lesioned voxels within each region of interest. In case of the NIHSS derivation cohort, patients with low WMH burden had a significantly higher frequency for how often the left insular cortex was affected (Fisher’s exact test: *p*=0.002, Bonferroni-corrected for multiple comparisons, all further parcels: *p*>0.05). Additionally, patients in this low WMH burden group had a higher lesion load in the left putamen (two-sided t-test *p*=0.01, Bonferroni-corrected for multiple comparisons, all further parcels: *p*>0.05).

## Discussion

Facilitated by our novel generative hierarchical modeling framework, we here, for the first time, investigated whether a pre-existing WMH burden is associated with stroke outcomes depending on specific stroke lesion patterns. We present evidence that, indeed, a higher level of pre-stroke WMH burden is linked to an aggravation of acute stroke severity in *lesion pattern-specific* ways. That is, WMH burden was not linked to a higher acute stroke severity in general, but was associated with a higher stroke severity in the case of specific lesion topographies only. More specifically, we observed that lesions relating to left-hemispheric insular and inferior frontal brain regions, as well as lesions affecting right-hemispheric temporo-parietal brain regions were associated with a higher stroke severity explicitly in those patients with a high WMH burden compared to those with a low WMH burden. This lesion-specific effect did not hold for long-term functional stroke outcomes: rather, the overall WMH burden was associated with higher odds of unfavorable outcomes across all strokes.

All in all, these findings suggest that an exacerbated disruption of network integrity by the combination of WMH and specific stroke lesions may be associated with a higher stroke severity and should be explored further in functional imaging studies that simultaneously considered information from both lesion types. Furthermore, our findings may be of particular clinical relevance, given that they motivate a lesion pattern-specific consideration of WMH burden in acute ischemic stroke, and, especially if confirmed in prospective studies, prompt tailored clinical management. For example, our findings elicit the testable hypothesis that high WMH burden patients may benefit from a more aggressive acute treatment and intense rehabilitation in case they experience left-hemispheric lesions in proximity of the insula or right-hemispheric temporo-parietal regions.

### Stroke severity – left-hemispheric lesion pattern

The involvement of the left inferior frontal gyrus suggests a potentially detrimental effect of global WMH burden on (Broca’s area-mediated) language function. This assumption of language-specific WMH effects is further supported by a wealth of stroke studies reporting associations of WMH with worse language outcomes,^19,20^ declines in language performance,^21^ and worse language recovery after stroke.^22^ Previous studies have shown that WMH predominantly impairs the function of long-range (rather than medium- or short-range) fibers.^19,35^ The restriction of physiological long-range fiber function, in turn, appears to mediate negative WMH effects on language outcomes, as a study by Wilmskoetter and colleagues suggests.^19^ Furthermore, intact language function is thought to depend on the integrity of both local, task-specific, as well as distributed domain-general networks.^36,37^ Hence, language function after direct focal lesions of language areas, such as Broca’s area, may be more excessively impaired in the case of concurrent diffuse pre-stroke brain pathology, such as WMHs affecting long-range fibers and by these means the integrity of spatially distributed brain networks.^38^

Overall, our findings motivate the more precise evaluation of WMH burden for patients presenting with left-hemispheric lesions in language areas. We here only assessed rather coarse-grained behavioral outcomes – with the additional limitation that the NIHS Scale is known to result in higher scores for left-hemispheric lesions.^39^ Future studies could therefore specify the impact of WMH and stroke lesion interaction effects on language outcomes or individual NIHSS items as a first step,^40^ corroborate the detrimental effects and estimate their clinical relevance further. Conceivably, a more consistent consideration of the WMH burden in models of language-related outcomes may substantially increase the prediction performance.

### Stroke severity – right-hemispheric lesion pattern

The WMH-modulated right-hemispheric lesion pattern detected in our analysis most prominently comprised inferior parietal lobule (IPL) brain regions, e.g., supramarginal and angular gyrus. These brain regions are known to fulfill a rich variety of cognitive functions^41,42,43^ especially visuospatial attention.^44^ Correspondingly, lesions of right IPL are frequently observed to lead to the clinical phenomenon of hemispatial neglect, i.e., the inability to orient attention to the contralesional body side.^45^ Intriguingly, Bahrainwala and colleagues previously demonstrated increasing odds of having neglect and having more severe neglect with increasing WMH burden, independent of stroke lesion volume, age, sex and race.^46^ Much like language, spatial attention may heavily rely on the integrity of large-scale (in this case fronto-parietal) brain networks,^47^ beyond cortical localization. This dependence on large-scale brain networks may underscore the importance of WM tracts connecting critical cortical brain regions. WMHs affecting these WM tracts may then compromise the integrity of these large-scale networks, augmenting the detrimental effect of locally specific (cortical) stroke lesions.

In analogy to our considerations in the previous paragraph on language outcomes, we see the same benefit of integrating WMH burden in prediction of attention deficits after right-hemispheric temporo-parietal lesions.

### Chronic functional outcomes

In our study, we did not observe any reliable lesion pattern-specific impact of WMH burden on mRS-defined functional outcomes. Instead, the *total* amount of WMH burden was associated with higher odds of unfavorable outcomes. This constellation of effects consequently stands in opposition to our stroke severity-specific findings, as we here uncovered lesion pattern-specific, but no overarching total WMH burden effect. Importantly, the effect of the continuous total WMH burden on functional outcomes was discernible despite accounting for clinical factors, such as age, sex, comorbidities, and stroke lesion pattern, as well as total stroke lesion volume. Two considerations may contribute to the explanation of these findings: Firstly, the mRS is even more coarse grained than the NIHSS. Specific symptoms post-stroke, such as aphasia and neglect, may thus not have been represented well enough in the mRS to have a comparable effect to the NIHSS analyses (as the NIHSS might be more sensitive to these deficits).^48^ Secondly, the process of neurorecovery may, like instantaneous language and attention performances, precisely rely on the integrity of large-scale networks connected through WM tracts and thus be negatively affected by WMHs.

### Limitations

Several additional limitations should be considered: The exact time points of outcome score acquisition were rather variable. We here strove to maximize sample size, and therefore accepted stroke severity scores that were obtained during the acute hospital stay and mRS scores recorded between 60 to 190 days post-stroke. Moreover, neither the pre-stroke functional status, nor the administration of acute stroke treatments, such as thrombolysis or mechanical thrombectomy, were systematically recorded in our cohort. Given that our data was mostly obtained prior to 2011, expected numbers of treated patients are low. Nonetheless, it may be of value to adjust for these additional clinical and also pre-stroke factors and harmonize time points of data acquisition in future studies. Our study cohort was slightly younger and more mildly affected than expectable for an unselected stroke patient cohort.^49^ This circumstance may be due to a more frequent failure to obtain informed consent from older and very severely affected patients. Nonetheless, we expect that the inclusion of these older and more severely affected patients would have enhanced, rather than decreased the here observed differences between patients with low and high WMH burden. Future studies are warranted to assess and quantify this assumption. We here focused on interpreting interaction effects of stroke lesion pattern and WMH burden. While we corrected for important covariates, such as a patient’s age, sex and comorbidities, it was beyond the scope of this study to present an exhaustive evaluation of (main and interaction) effects of these factors. These aspects will be addressed in future work. Furthermore, we here relied on scans, as obtained in clinical routines in multiple countries and clinical sites, with associated challenges arising through data heterogeneity. However, both the automatic stroke and WMH lesion segmentations comprised thorough quality control steps aiming to ensure a high quality of individual lesion segmentations.^25,26^ In addition, our entire pipeline, especially the NMF-derived lesion pattern representation, could be easily transferred to completely new data. We did not account for the actual stroke lesion when computing the WMH burden. Of note, most patients in our study featured rather small lesions though, with a median size of 3.1ml. Altogether, we thus do not estimate our results to be decidedly altered if stroke lesions were included in WMH estimation. Finally, it may be fruitful to incorporate precise WMH lesion locations. Conceivably, the disruption of specific WM tracts may be particularly relevant, being intimately linked to specific functions. Moreover, as it was beyond the scope of the current study, we neither included information on further markers of small vessel disease, such as microbleeds or lacunar infarcts,^7,8^ nor did we incorporate measures of pathological changes of normal-appearing white matter, such as the peak width of skeletonized mean diffusivity (PSMD).^50^

## Conclusions

In this study, we introduced a novel modeling framework to investigate WMH-specific stroke lesion pattern effects, with WMH burden being a marker of pre-stroke brain health. With respect to acute stroke severity, we observed that lesions of left-hemispheric insular and inferior frontal brain regions and right-hemispheric temporo-parietal brain regions were associated with substantially higher scores in case of high WMH burden. The spatial distribution of these lesion patterns implicates two of the most fundamental cognitive functions, language and spatial attention. On the other hand, modified Rankin Scale-defined functional outcomes ∼3 months after stroke were influenced by the global WMH burden, rather than specific lesion patterns. Our results permit novel insights into WMH effects: especially if confirmed in prospective studies, our findings may spur lesion pattern- and WMH burden-specific modifications of acute treatments and long-term rehabilitative approaches.

## Data Availability

The authors agree to make the data of development and validation cohorts available to any researcher for the express purposes of reproducing the here presented results, and with the explicit permission for data sharing by Massachusetts General Hospitals institutional review board.

## Abbreviations

AF: atrial fibrillation
AUC: area under the curve
CAD: coronary artery disease
DM: diabetes mellitus
HTN: hypertension
NIHSS: National Institutes of Health Stroke Scale
NMF: non-negative matrix factorization
mRS: modified Rankin Scale
PCA: posterior cerebral artery
WMH: white matter hyperintensities

## Acknowledgments

We are grateful to our colleagues at the J. Philip Kistler Stroke Research Center for valuable support and discussions. Furthermore, we are grateful to our research participants without whom this work would not have been possible.

## Author contributions

A.K.B. and N.S.R. conceived and designed the study, led data interpretation, and prepared the manuscript. A.K.B. led data analysis, M.B., S.H., M.D.S., R.W.R., M.B., O.W., D.B. also contributed to data analysis. E.M.A., K.L.D., M.J.N., A.V.D., A.K.G., M.R.E., B.L.H., B.J.T.M., E.C.M., J.A., O.S.B., S.B., J.W.C., A.D., C.J.G., L.H., L.H., K.J., J.J.C., S.J.K., R.L., C.R.L., C.W.M., J.F.M., C.L.P., A.R., S.R., J.R., J.R., T.R., R.L.S., R.S., P.S., A.S., M.S., A.S., T.M.S, D.S., T.T., V.T., A.V., J.W., D.W., R.Z., P.F.M., B.B.W., C.J., A.G.L., J.M., P.G., O.W. contributed to data acquisition, management and preprocessing. All authors contributed to results interpretation and final manuscript preparation.

## Supplemental materials

### Methods MRI-GENIE

The MRI-Genetics Interface Exploration (MRI-GENIE) study is a large collaboration of 12 international sites that assembled sociodemographic/clinical, neuroimaging and genotypic data from 3,301 patient. MRI-GENIE’s main aim was the genetic analysis of acute and chronic cerebrovascular neuroimaging phenotypes as extractable from clinical-grade MRIs of acute ischemic stroke patients.^1^ The study furthermore put a prime on the availability of Causative Classification of Stroke (CCS). Six sites could contribute acute stroke severity and ∼3 months functional outcome data and are considered in the present study. More specifically, these six studies were: BASICMAR (Spain, 124 patients in total), SAHLSIS (Sweden, 401 patients), LUND STROKE REGISTER (Sweden, 196 patients), LEUVEN (Belgium, 448 patients), GCNKSS (USA, 245 patients) and GASROS (USA, 457 patients; c.f., ^1^ for further study characteristics, such as mean age and percentage of men/women).

### Sample size calculation

Out of 2,765 automatically segmented stroke lesions,^2^ 1,920 (70.1%) passed internal quality control by two experienced raters (M.B., A.K.B.). The total number of finally included patients was determined by the additional availability of NIHSS/mRS outcomes (sample size for the lesion representation derivation: n=1,107 patients) and the availability of WMH lesion segmentations (resulting in n=928 patients for NIHSS, and n=698 patients for mRS analyses with largely overlapping samples).

### Information on Neuroimaging parameters

All images were either obtained on 1T, 1.5T or 3T scanners (General Electric Medical Systems, Philips Medical Systems, Siemens, Toshiba, Marconi Medical Systems, Picker International, Inc.).

### Axial T2-Fluid-attenuated inversion recovery (FLAIR)-weighted images

The mean in-plane resolution had a mean of 0.7 mm (minimum – maximum: 0.3-1.0mm), the mean through-plane resolution was 6.2mm (3.0-30.0mm)

### Diffusion-weighted images (DWI)

Axial scans were obtained in the majority of cases (2727/2770 axial, 43/2770 coronal). Axial: The reconstruction matrix was mostly 256×256mm^2^, but ranged from ranged from 128×128mm^2^ to 432×384mm^2^. The median field-of-view totaled 230 mm and ranged from 200-420 mm. Slice thickness had a median of 5mm and ranged from 2-7mm with gaps of 0-3mm. Median TR was 4.773ms, median TE 92ms. Coronal: median TR 8.200ms and TE 112ms, 5mm thickness, field-of-view 260mm, reconstruction matrix 256×256 mm^2^. Sequences were mostly made up of 3 directions, but the range was from 3-25. Most scans had one low b-value=0s/mm2 (range 0-50s/mm2) and one high b-value of 1000s/mm2 (range 800-2000s/mm2).

### Model specification for stroke severity

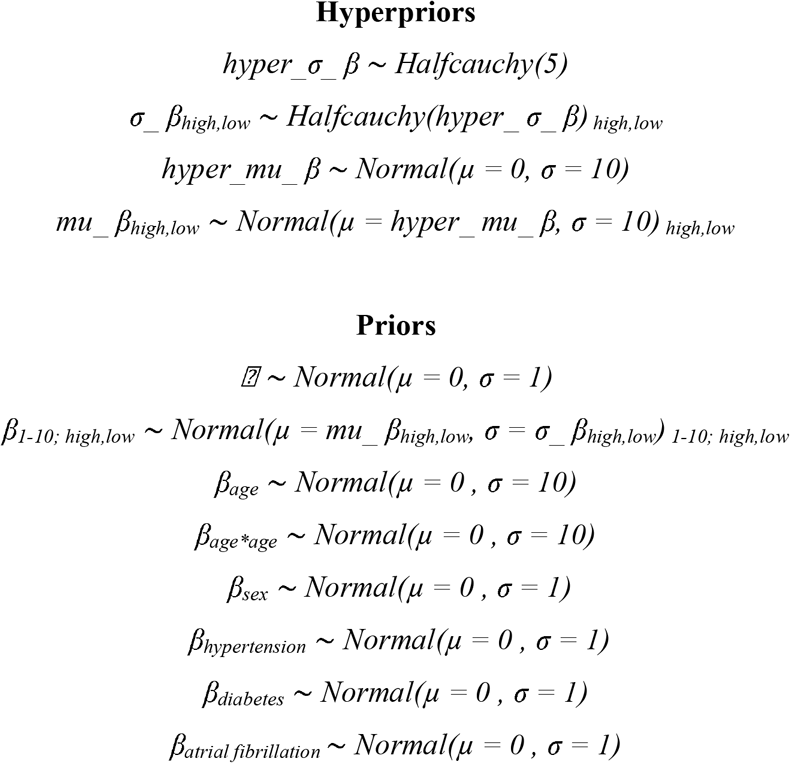

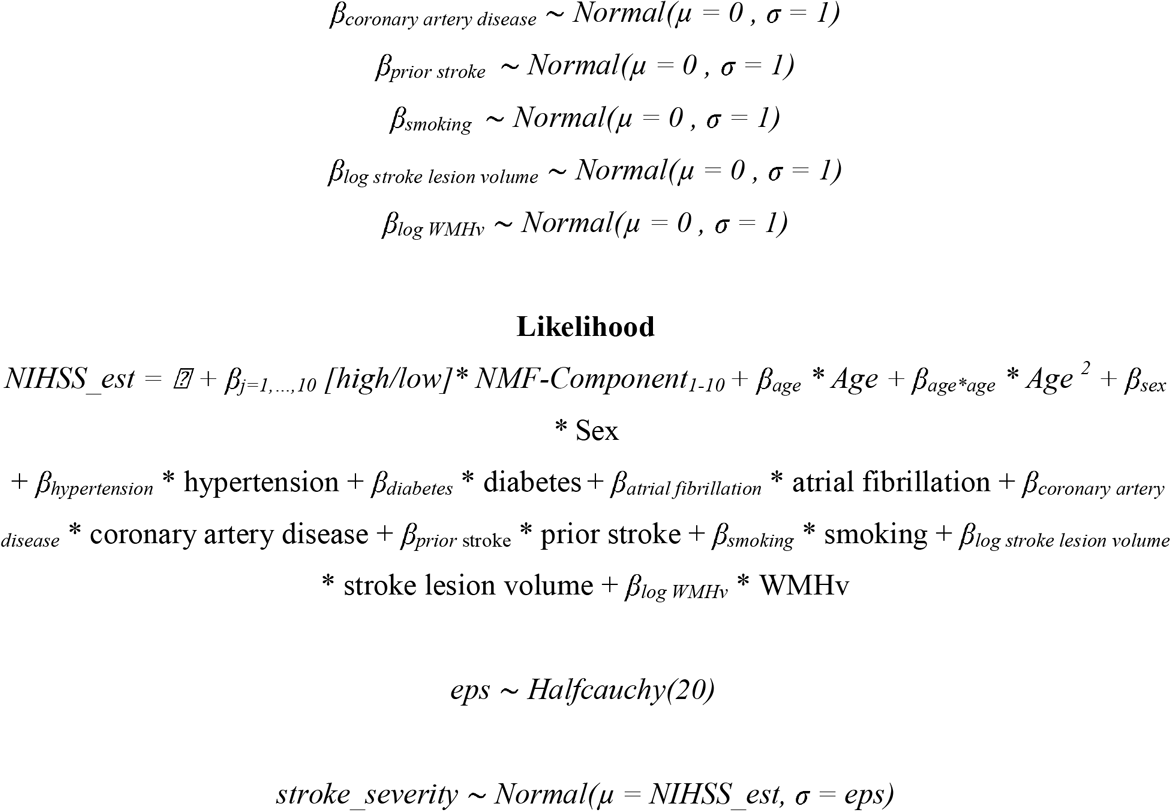

### Model specification for the long-term functional outcomes

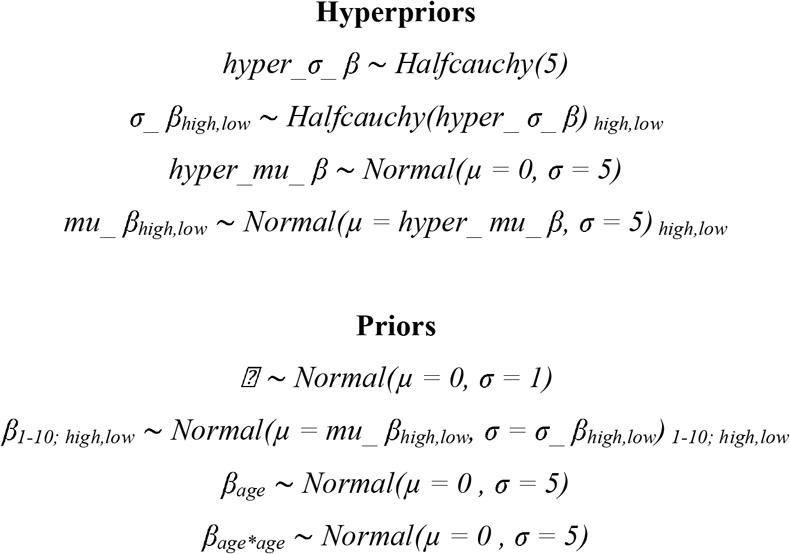

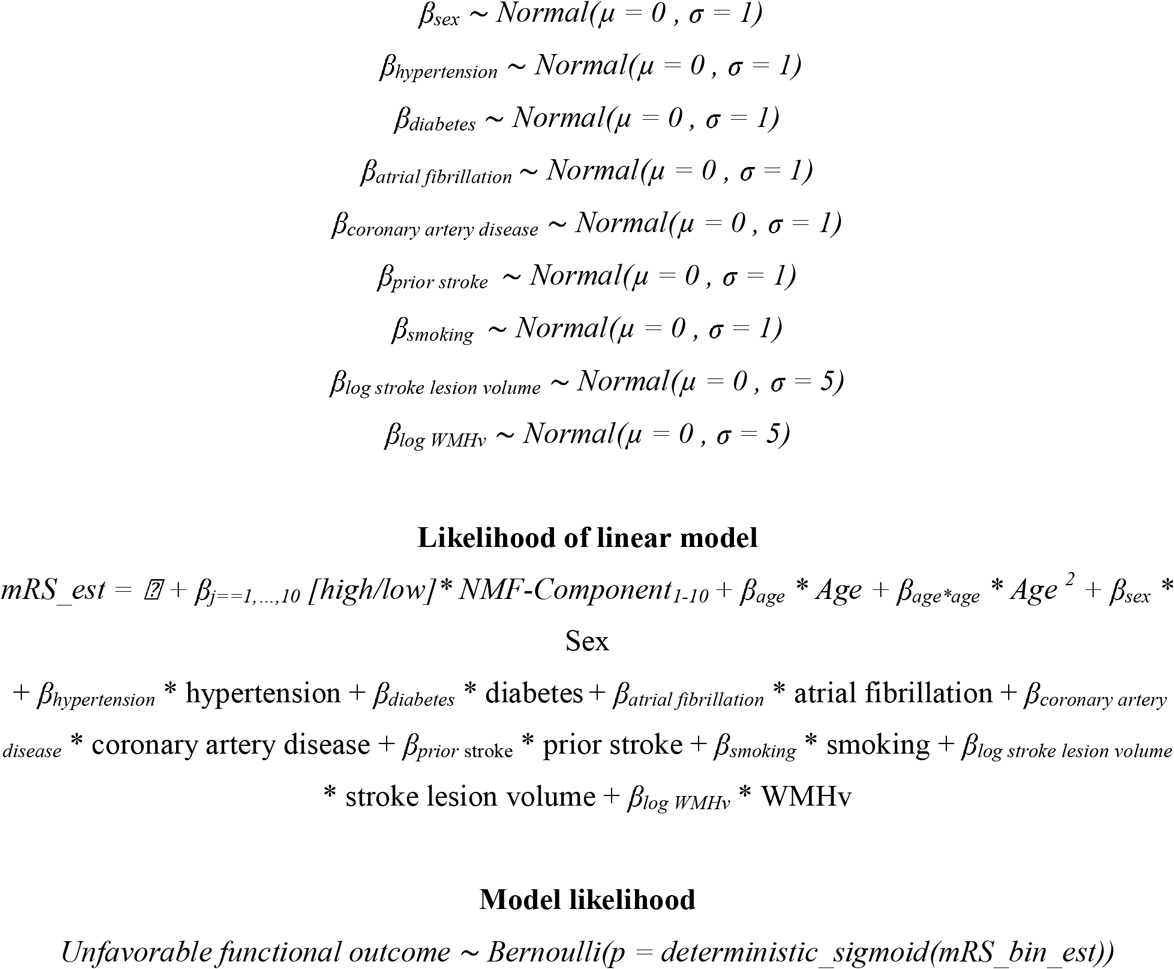

### Sampling from the posterior

We determined the posterior distributions of regression parameters by employing the No U-Turn Sampler (NUTS), a type of Monte Carlo Markov Chain algorithm (setting: draws=5000).^5^

### WMH burden-specific lesion pattern effects

We investigated differences in lesion pattern effects in patients with high and low WMH burden by subtracting their posterior distributions. We assumed a substantially altered lesion pattern effect between patients with varying amounts of WMH burden in case of difference distributions not overlapping with zero. We adjudicated on a substantial overlap with zero based on the 90% highest probability density interval (HPDI), as already introduced in our previous work.^3^

## Results

### Explaining acute stroke severity: Validation

As in the derivation cohort analysis, bilateral subcortical brain regions and white matter tracts had the highest relevance in explaining stroke severity across both groups of patients with high and low WMH burden (**Supplementary Figure 1A**). Furthermore, we extracted similar patterns of differences between high and low WMH burden groups: *Lesion patterns* 4 and 6 had a substantially higher relevance for high WMH burden than low burden patients (*lesion pattern* 4: mean difference= -0.8, 90%-HPDI= -1.5 to -0.1; *lesion pattern* 6: mean difference= -1.9, 90%-HPDI= -3.2 to -0.4). Additionally, there was one extra lesion pattern, *lesion pattern* 10, combining left-hemispheric brain regions in posterior cerebral artery (PCA)-territory, that was more relevant in high versus low WMH burden patients (*lesion pattern*10: mean difference= -0.8, 90%-HPDI= -1.5 to -0.1).

### Explaining unfavorable functional outcomes: Validation

When re-fitting the Bayesian model in the validation cohort, the most relevant brain regions were subcortical regions in both hemispheres, as well as cortical regions in the left hemisphere **(Supplementary Figure 1B)**. There was no observable effect for the overall continuous amount of WMH burden (mean=-0.01, 90%-HPDI=-0.2 – 0.7), yet the relevance of *lesion pattern* 1 was substantially higher for the high WMH burden group (mean difference=-0.7, 90%-HPDI=-1.2 - - 0.1). Nonetheless, no individual lesion pattern effect for high and low WMH burden groups diverged considerably from zero.

**Supplementary Figure 1.**
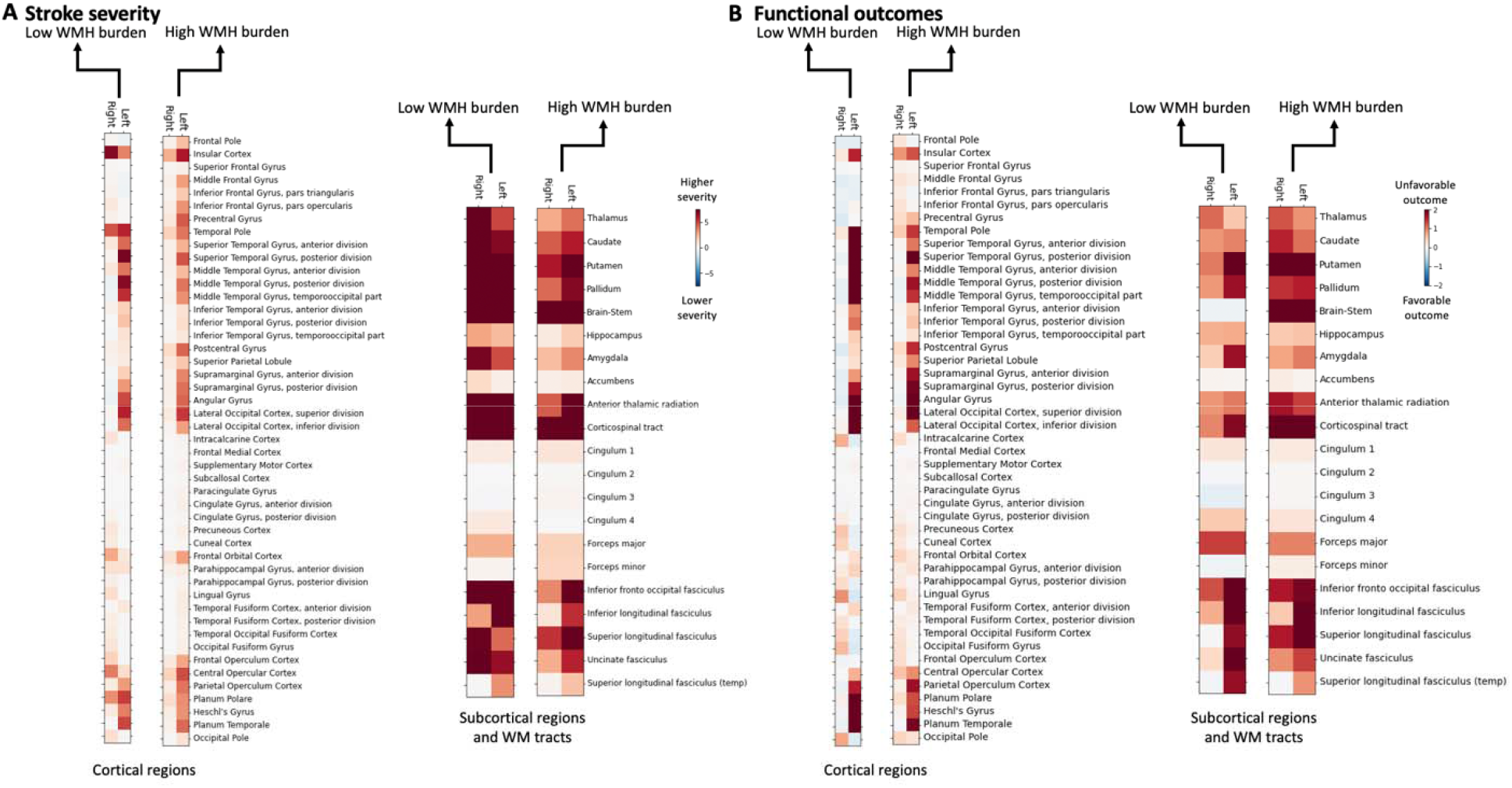
Brain region-wise relevances for explaining stroke severity (A) and functional outcomes (B) in the validation cohort. Similar to the relevances estimated in the derivation cohort, subcortical grey matter brain regions and white matter tracts of both hemispheres were linked to both, higher stroke severity and higher odds of unfavorable outcomes. Additionally, lesions in cortical brain regions, comprising superior and middle temporal gyrus, Herschl’s, supramarginal and angular gyrus and lateral occipital cortex, mainly in the left hemisphere, contributed to a higher stroke severity and unfavorable outcomes. These observations were the same independent of a high or low WMH burden status.

